# Mental wellbeing in the Bangladeshi healthy population during nationwide lockdown over COVID-19: an online cross-sectional survey

**DOI:** 10.1101/2020.05.14.20102210

**Authors:** Mohammad Ali, Gias U. Ahsan, Hasinur R. Khan, Ahmed Hossain

## Abstract

**Background:** We aim to profile and contextualize the level of positive mental health among the healthy population during the beginning of nationwide lockdown over COVID-19 in Bangladesh.

**Design and setting:** An online sample of 1404 healthy individuals was collected through the authors’ networks with residents and popular media in Bangladesh. The survey was conducted between March 27, 2020, and April 7, 2020, following the Bangladesh government’s lockdown announcement.

**Method:** A questionnaire comprising the Warwick Edinburgh Mental Wellbeing Scale (WEMWBS) and sociodemographic information was posted to an online survey.

**Results:** The mean wellbeing value was 38.4 (standard deviation, 11.2), indicating a lower mental health level exist in adults. Also, the mean wellbeing score for males was 39.0 (10.8) units compared to females with scores 37.0 (11.2), and the highest scores for government workers was 41.2 (11.8). Unemployment (35.6) or business employees (35.5) have a lower level of mental health. In the lockout days, the elderly population of age 50 years and above had high day-to-day variation of wellbeing scores. After confounding adjustment in multivariable linear regression models, there found a better wellbeing scores for males (estimate = 1.79, 95% CI = 0.5, 3.1), and government-workers (estimate = 5.86, 95% CI = 2.2, 9.5). Moreover, the never-married female had significantly higher well-being score compared to married women (estimate = 3.31, 95% CI = 1.0, 5.7).

**Conclusion:** The COVID-19 pandemic has been associated with low mental wellbeing that indicates depression in our study. Suggestions for improvement will be implemented to promote the mental health to women, unemployed and business communities. Older people 50 years of age and over reported a greater day-to-day variation in their mental health. The married women with their decreased mental wellbeing should be given special consideration. The research will help let clinicians and policymakers decide where the measures can be implemented to improve their mental health during and after this pandemic.

## Background

Novel coronavirus disease (COVID-19), a viral infection caused by SARS COV-2, spread in December 2019 from Wuhan, China and declared a pandemic in March 2020 [1]. Rapid human-to-human transmission of this virus has been widely confirmed, with no clinically approved antiviral medication or vaccine to be used for treatment or prevention [2]. Social-distancing and home quarantine recommended by the World Health Organization (WHO) has been proven to be an effective way to minimize the speed of this virus spread. Thus governments of many countries have locked down their countries due to COVID-19. During the lockdown period, all the business centres, government, and non-government offices are closed. Restrictions also apply to any social gathering after closing all transport modes which forced citizens to be isolated or quarantined at home [3].

Fear of being infected with COVID-19 and uncertainty due to lock down have an immense psychological negative effect on the general population [4]. In fact, social quarantine also affects human psychology and mental health [5]. A study showed that there is a strong connection between quarantine and cognitive function, neuroendocrine and cardiovascular system that can cause depression, cognitive impairment and sleep disturbance [6].Some studies have found that quarantine can cause adverse behavioral patterns including substance abuse, crime and even suicide [7–8]. A suicide case relating to quarantine has been reported in Bangladesh due to COVID-19 [9]. A recent review also indicated that negative psychological effects such as symptoms of post-traumatic stress, frustration and rage along with long-lasting quarantine effects are widespread among quarantined populations [10]. Thus mental well-being data during the lockdown period is needed to assist the policymaker in assessing this pandemic’s current and post-pandemic situation.

Research has indicated a close link between poor mental health and financial insolvency and instability [11]. It was predicted that record 3.3 million people could only lose their jobs in the US during and after this pandemic. The situation could be worse in low-income countries like Bangladesh than the high-income countries [12]. Therefore the psychological problems relating to economic instability are an apparent consequence of people being locked down.

Positive mental health or mental well-being has recently emerged as an important predictor of overall health and longevity [7–9]. Mental well-being is more than the absence of mental illness or psychiatric pathology. It implies ‘feeling good’ and ‘functioning well’ and includes aspects such as optimism, happiness, self-esteem, resilience, agency autonmy and good relationships with others [7–11,13–14]. Mental wellbeing has also been found to be associated with better occupational functioning in adulthood [1–2,15].

Pathophysiology, epidemiology and clinical management of COVID-19 are the main research subject during this pandemic [16]. The psychological effects of such a pandemic could nevertheless be left to millions of people with a long-term or irreversible psychiatric disorder with which negative social implications could begin. Preventing emotional and behavioral issues and encouraging physical and social and mental well-being should therefore be a priority for the people of Bangladesh. The research would further identify those individuals who need support that can help strengthen the mental health care mechanisms. Thus the study aims at measuring the mental health of Bangladesh’s general population during the COVID-19 pandemic lockdown phase.

## Method

### Study population

In this representative sample of the healthy adult population of Bangladesh, we tried to measure cross-sectional associations between the sociodemographic factors and mental wellbeing. On 26 March 2020, Bangladesh called for an active lockdown. Bangladeshi citizens were invited to participate in a study via social media during the onset of the lockdown time. Participating in the study we used Facebook, Email, Whatsapp, Imo, and Skype. Approximately 7000 people received the invitation from March 28 to April 7, 2020. Of a potential total of 7000 online connections, 1483 participated, giving 21.2% overall response rate. Of these, 1404 responses were considered for analysis after consideraiton of the inclusion and exclusion criteria. The participant must be above 16 years of age and resident of Bangladesh residing either in the town or in the village. Participants with missing values were excluded, to avoid bias.

### The questionnaire

The online questionnaire, produced in Survey Monkey, included a wide variety of demographic variables such as gender, age, current living place, marital status, occupation, and education status. The age group is categorized as: 16–19, 20–29, 30– 39, 40–49, > = 50. Current working conditions were also taken over during the lockdown period and are categorized as: not working, working from home, working both at home and outside.

### Warwick Edinburgh Mental Wellbeing Scale

The Warwick-Edinburgh Mental Well-being Scale (WEMWBS) was developed to provide an appropriate measure of mental well-being for use in adults. WEMWBS was found to be easy to complete, clear and unambiguous in research carried out with adults and proved popular with both practitioners and policy makers [12,16]. The WEMWBS measures subjectively perceived wellness and psychological function on a 5-point Likert scale over the last 2-week period (14–70 points, higher = better mental health). The scale was not invented as a mental illness screening method and there is no cut off point [10]. WEMWBS was used in various studies and validated in several languages and settings [11,13–14]. A user licence was received to include WEMWBS in the study (Regitration ID: 517150559).

### Statistical Analysis

Descriptive statistics (including means and standard deviations) were calculated for WEMWBS scores. ANOVA was used to test for an association between sociodemographic factors and mental wellbeing scores (WEMWBS). In multivariable linear regression models, associations between WEMWBS and socio-demographic variables were investigated, with WEMWBS as the dependent variable, and sociodemographic variables as independent variables. Factors at the 5% level were defined as statistically significant. We evaluated the internal consistency of WEMWBS using Cronbach’s alpha [17].

## Results

### Socio-demographic characteristics

Table 1 shows the sociodemographic characteristics in comparison to the WEMWBS ratings. Of the 1404 respondents, 63.2% and 36.8% were male and female, respectively. Participants were overwhelmingly educated, with 83 percent being undergraduates or above, meaning they lived in lower-middle, middle or higher affluent households. The mean score for WEMWBS was 38.5 (standard deviation, SD 11.1) and the median was 38. During the COVID-19 pandemic, male scores were 1.7 points higher than for female suggesting a lower mental health level among female participants. Cronbach’s alpha for WEMWBS was 0.88 (95% CI [0.85; 0.90]) which indicates an internal consistency of the WEMWBS ratings. A significant association was found between WEMWBS scores and occupation. More variabilities are observed in age groups 16–19 and older than 50 years which indicates these groups of participants were psychologically unstable.

**Table 1:** Participant characteristics and association with wellbeing scores.

| Variables | Categories | n | Mean (sd) | P-value* |
| --- | --- | --- | --- | --- |
| Age group | 16-19 | 50 (3.6%) | 38.7 (12.78) | 0.087 |
|  | 20-29 | 767 (54.6%) | 38.06 (10.85) |  |
|  | 30-39 | 447 (31.8%) | 38.24 (10.35) |  |
|  | 40-49 | 110 (7.8%) | 41.12 (10.89) |  |
|  | >=50 | 30 (2.1%) | 40.07 (12.65) |  |
| Gender* | Male | 888 (63.2%) | 37.37 (10.82) | **0.007** |
|  | Female | 516 (36.8%) | 39.04 (11.22) |  |
| Marital Status | Married | 713 (50.8%) | 38.29 (10.97) | 0.799 |
|  | Never-Married | 671 (47.8) | 38.60 (10.91) |  |
|  | Others | 20 (1.4%) | 37.40 (12.63) |  |
| Education | Schooling 6-12 years | 232 (16.5%) | 38.03 (11.18) | 0.777 |
|  | Undergraduate | 576 (41.0%) | 38.37 (10.83) |  |
|  | Graduate | 596 (42.5%) | 38.63 (11.18) |  |
| Occupation | Business | 79 (5.6%) | 35.53 (11.42) | <**0.001** |
|  | Government | 69 (4.9%) | 41.16 (11.79) |  |
|  | Healthcare | 356 (25.4%) | 39.62 (09.99) |  |
|  | Housewife/ Unemployed | 129 (9.2%) | 35.57 (10.40) |  |
|  | Non-government | 353 (25.1%) | 38.77 (11.66) |  |
|  | Student | 418 (29.80) | 38.10 (11.31) |  |
| Working Condition | Not employed | 758 (54.0%) | 37.94 (11.12) | 0.201 |
|  | Work from home | 422 (30.1%) | 38.93 (11.09) |  |
|  | Work from both home and Outside | 224 (15.9%) | 39.13 (11.01) |  |
| Current location of living* | City | 1118 (79.1%) | 38.68 (11.09) | 0.0961 |
|  | Village | 286 (20.9%) | 37.45 (11.12) |  |
| *p-value was calculated from ANOVA, and the p-values for gender and current location of living were calculated from t-test. |  |  |  |  |

**Figure 1:**
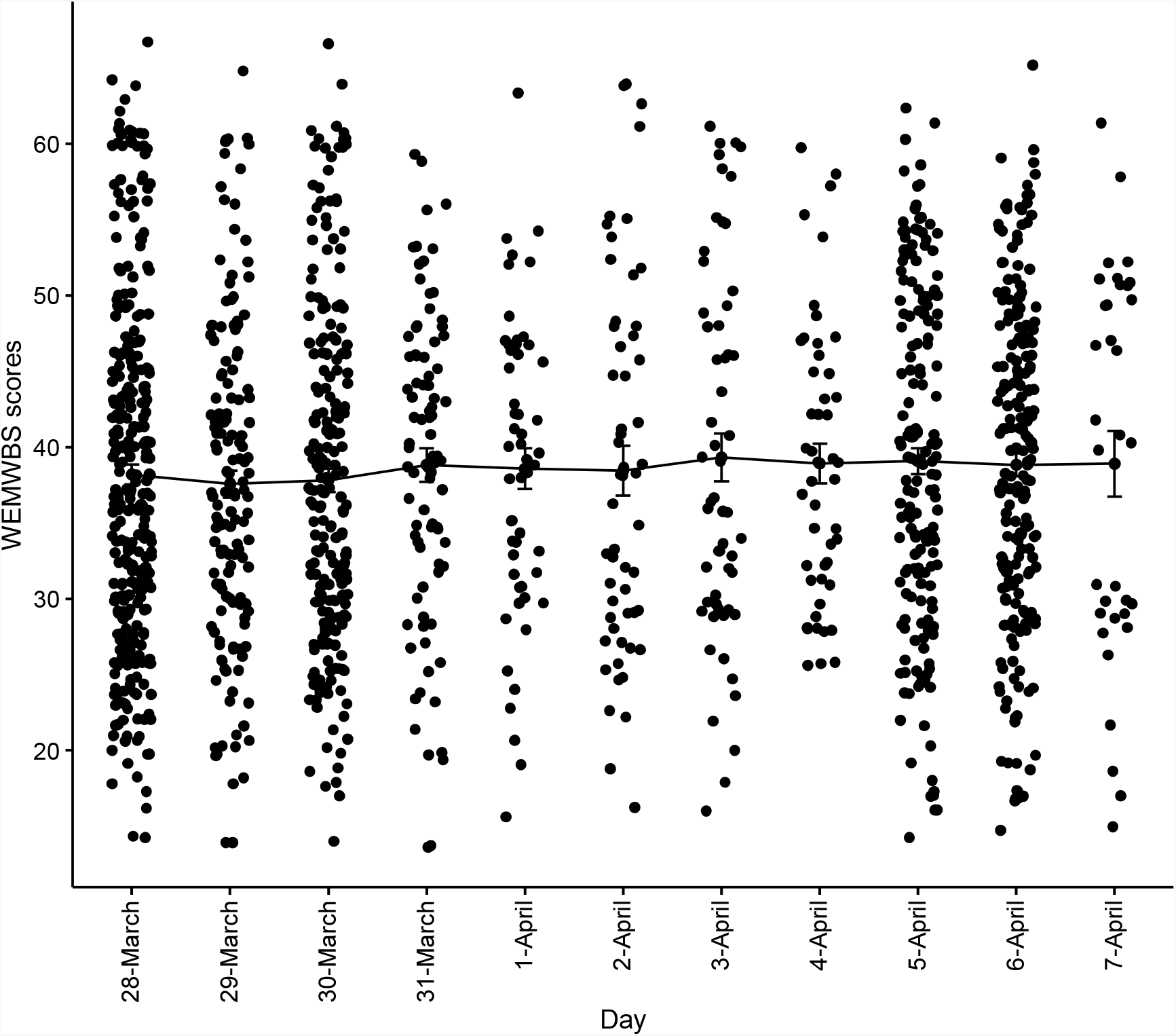
Day to day change of mean wellbeing scores during lockdown period.

**Figure 2:**
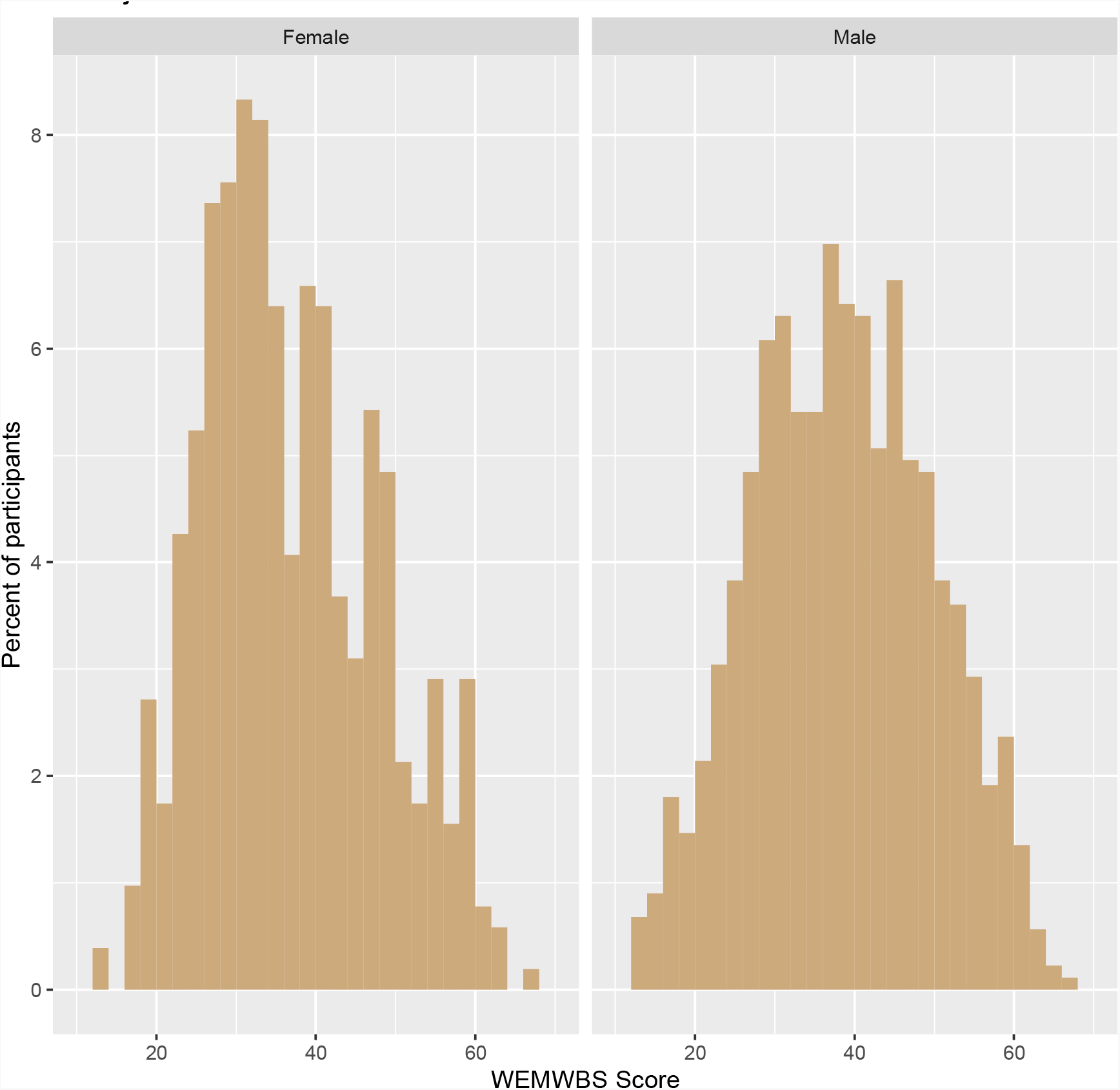
Histogram of unadjusted mental wellbeing scores by male and female.

The WEMWBS scores ranged from 14 to 67 (see Figure 1), i.e. the range of possible values from 14 to 67 was used. It appears from the Figure 1 that the mean wellbeing scores is nearly 38 for March 7, 2020 and did not change as the days changed. It suggests a lower mental health level during the COVID-19 pandemic. However, the rising number of COVID-19 cases per day in Bangladesh since the start of the lockout period had no significant effect on the mental wellbeing indicates the current lower mental health wellbeing started early due to global stressful situations. In addition, the Figure 2 shows that scores are almost normally distributed for the male group. But the scores suggest higher variability for the female community. It indicates in the lockdown time female was mentally disturbed.

### Items of WEMWBS scores

Figure 3 shows WEMWBS with items numbered, each of which was presented as percentage in turn by participants. Most participants reported negatively on Items 3 (“feeling relaxed”) and 4 (“interested in other people”) during the COVID-19 time. Looking at sub-scales of WEMWBS, it was found 75% of all participants were ‘rarely’ or ‘none of the time’ interested in other people, and about 60% of participants did not feel relax (item 3) during this pandemic time. In addition, at this time almost 60 percent of the participants did not feel confident (item 10). On the contrary, about 50% of the participants can feel they are useful (item 2) and can make up own mind about things (item 11).

**Figure 3:**
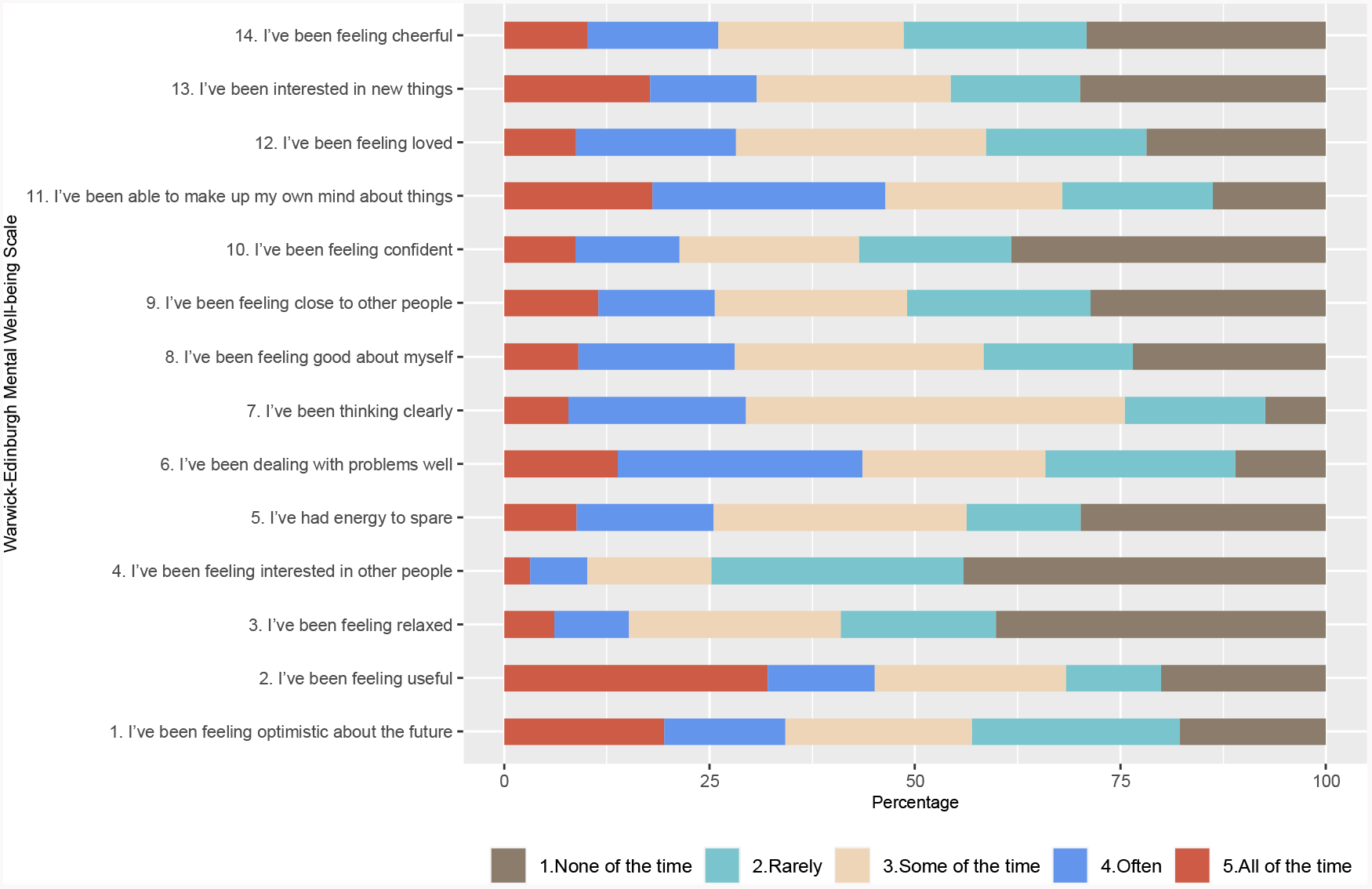
Percentage of participants by the items of WEMWBS.

### Trajectories of scores by Socio-demographic factors

The Figure 4 shows a comparison of day to day variation by different categories of sociodemographic factors since the start of the survey at the beginning of the lockdown period. The rising scores seem a similar trend for both male and female as day changes. Compared to other age groups, the variability of scores in age group 50 and above is much higher indicates that the participants from this age group deviated that scores as the day changes. It indicates that the elderly population may be more disturbed by the pandemic situation. Similar types of instability have been observed in the divorced / separated population and in the people living in the village during survey time. The participants who were housewives or unemployed are in the non-working group and found WEMWBS scoring low but the variability is constant while the participants who work at home and work both at home and outside of home were found to be inconsistent with the changing days.

**Figure 4:**
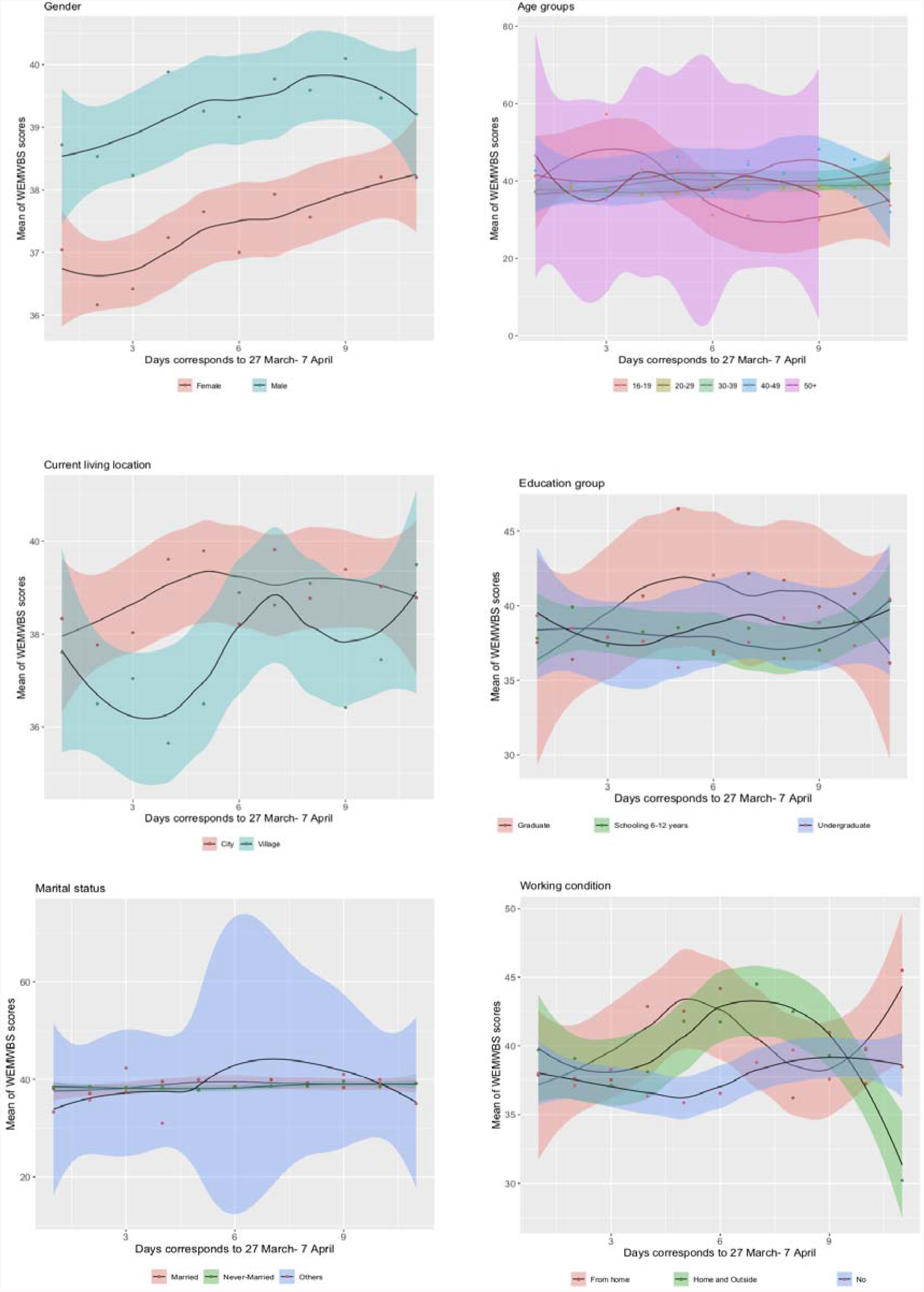
Day to day mean wellbeing scores by sociodemographic variables.

### Multivariable Linear Regression Models

Table 2 presents the results from multivariable linear regression models. The results present the slope estimate, 95% confidence interval of the slope and the corresponding p-value. We applied 3 models. The first model is with all the data and the next two models were for male and female participants, respectively. We stratify the gender and applied the two models to avoid the confounding bias. It appears from the Model 1 that gender and occupational groups have had significant effect on the mental wellbeing scores. Male had a significantly higher feelings of wellbeing by an increment of 1.79 scores (slope estimate = 1.79, 95% CI = 0.5 to 3.1) than female. The participants who were involved in business had lower levels of wellbeing than government employees (by 5.87 units, p = 0.01) or health care (by 4.98, p = <0.001) or employees of private companies (by 3.31, p = 0.02). Model 2 provides the similar results because male was predominant in this study. Most women are not in the working group and therefore the occupational factor is not an important factor for the women participants. The unmarried female group participants appear to have higher well-being scores than the married women (by 3.31, p = 0.01).

**Table 2:** Results from multivariable linear regression model.

| Variable | Categories | Model 1 (All data) | Model 2 (Male data) | Model 3 (Female data) |
| --- | --- | --- | --- | --- |
| Age | 16-19 | -0.71, (-6.1, 4.7), 0.80 | 3.37, (-3.3, 10.1), 0.32 | -5.98, (-15.4, 3.5), 0.22 |
|  | 20-29 | -1.56, (-5.8, 2.7), 0.48 | 1.66, (-3.7, 7.1), 0.55 | -5.12, (-12.4, 2.2), 0.17 |
|  | 30-39 | -1.24, (-4.4, 2.9), 0.56 | 0.74, (-4.4, 5.9), 0.78 | -3.64, (-10.9, 3.6), 0.32 |
|  | 40-49 | 1.74, (-2.7, 6.2), 0.45 | 3.81, (-1.7, 9.3), 0.17 | -0.84, (-8.9, 7.2), 0.84 |
|  | >=50 | Reference |  |  |
| Gender | Male | **1.79, (0.5, 3.1), 0.01** | - | - |
|  | Female | Reference |  |  |
| Occupation | Government | **5.86, (2.2, 9.5), 0.01** | **7.88, (3.6, 12.1), <0.001** | -0.49, (-9.7, 8.8), 0.91 |
|  | Healthcare | **4.98, (2.2, 7.8), <0.001** | **4.17, (1.0, 7.3), 0.01** | 3.24, (-4.9, 11.4), 0.44 |
|  | Housewife/<br>Unemployed | 1.56, (-1.7, 4.8), 0.34 | 3.82, (-0.6, 8.3), 0.09 | -2.00, (-10.3, 6.3), 0.64 |
|  | Private | **3.31, (0.6, 6.0), 0.02** | **3.38, (0.5, 6.3), 0.02** | 0.98, (-7.4, 9.3), 0.82 |
|  | Student | **2.97, (0.1, 5.9), 0.04** | **4.10, (0.8, 7.4), 0.01** | -1.40, (-9.8, 7.0), 0.74 |
|  | Business | Reference |  |  |
| Working condition | Not-working | -0.84, (-2.2, 0.5), 0.22 | -1.29, (-3.0, 0.4), 0.14 | -0.21, (-2.4, 1.9), 0.85 |
|  | Both home and outside | -0.67, (-2.5, 1.2), 0.48 | -0.41, (-2.7, 1.8), 0.72 | -1.53, (-4.7, 1.7), 0.35 |
|  | Work from home | Reference |  |  |
| Marital status | Never married | 1.13, (-0.4, 2.7), 0.15 | -0.63, (-2.8, 1.5), 0.56 | **3.31, (1.0, 5.7), 0.01** |
|  | Divorced/<br>Separated | -0.02, (-5.0, 4.9), 0.99 | -7.44, (-17.5, 2.7), 0.15 | 2.30, (-3.4, 7.9), 0.42 |
|  | Married | Reference |  |  |
| Current living location | Village | -1.02, (-2.5, 0.5), 0.19 | -1.10, (-2.9, 0.7), 0.22 | -1.61, (-4.8, 1.5), 0.32 |
|  | City | Reference |  |  |

### Comparison with existing literature

During the lockdown time due to COVID-19 the general population sample in Bangladesh recorded average mental health score as 38.4. In Bangladesh, the mental health scores were much lower compared to the general population in Denmark (52.2) or the general population in England (49.8). Northern Ireland’s health care professionals (50.2) or Pakistan’s health care professionals (48.1) received much higher scores compared to health care professionals in Bangladesh (39.2). Thus, health care professionals in Bangladesh tended to have substantially lower mental wellbeing score during the pandemic period compared to any occupation in other countries. The worst WEMWBS score was seen in the inmates on remand from Scotland (37.1) and the rating is almost identical to Bangladesh’s wellness score.

**Table 3:** Comparison of wellbeing scores with existing literature.

| Country | Categories of participant | Year | WEMWBS Score (SD) | Time |
| --- | --- | --- | --- | --- |
| Bangladesh | General Population | 2020 | 38.4 (11.3) | During lockdown due to COVID-19 |
| Scotland [26] | Prisoners | 2017 | 41.2 (12.3) | Prison period |
| Scotland [26] | Prisoners | 2017 | 37.4 (12.0) | Remand period |
| UK [27] | Bereaved carers | 2019 | 46.4 (10.7) | Bereaved cares period |
| Denmark [28] | General population | 2016 | 52.2 (n/a) | Normal time |
| Spain [28] | General Population | 2016 | 58.1 (n/a) | Normal time |
| England [28] | General population | 2016 | 49.8 (n/a) | Normal time |
| Pakistan [29] | Healthcare worker | 2014 | 48.1 (9.4) | Normal time |
| China [30] | University student | 2018 | 47.4 (8.9) | Normal time |
| Brazil [31] | General population | 2014 | 51.8 (8.0) | Normal time |
| Norway [32] | primary health care patients | 2016 | 37.3 (12.6) | At hospital |
| Northern Ireland [33] | Health care professionals | 2017 | 50.2 (8) | Normal time |

### Discussion

This was the first study assessing Bangladeshi adult population mental health over different professional groups. Throughout the lockdown period, residents reported substantially lower mental health relative to citizens from other countries at a normal time. We used a Bangla version of WEMWBS to define mental wellbeing, and in this adult population group we found a strong internal consistency with a high Cronbach alpha.

Despite high levels of stress due to COVID-19 in Bangladesh, males had the highest level of mental health compared to females. Contrary to the study from Murray et al, in which women had significantly higher mental wellbeing than men, women’s mental well-being in the present sample was lower than men’s (+1.7 to – 1.7 units in this survey). Similar to our study, during the COVID-19 situation, a study among health care workers in China found female are more anxious than men [18]. Another research also found females in this pandemic situation were in the lower mental wellbeing [19].

Our results reflect more day-to-day variation in age group 50 and over mental wellbeing. Findings at the population level to which the scale is intended are stable.

Working in a government position has been found to be associated with higher mental health (increase of 1.6 units) compared with the people involved in business. One explanation for that could be business financial uncertainty. A retrospective research indicates a strong link between financial solvency and good mental wellbeing [20]. Similar to our results, a report found that business and career uncertainty have a substantial negative effect on the mental health of adult population [21].

Although being single was associated with increased wellbeing for female participants (increase of 2.0 units) compared to married women. Compared to regular time, married women can face a burden with a high household workload, which results in less private time. The arrangements for household work may have increased to meet the needs of their families, and children. One research concluded that increased household load raises common mental illness [22]. These results agree with findings from other studies which defined the pandemic period as the most distress and difficulty characterized phase [23–25].

It is accepted that the research had limitations: First, we were unable to include dietary habit, physical activity in these analyzes because no data were collected on this behaviour. Second, while the study’s cross-sectional nature helped us to explore the associations between mental well-being and multiple behaviours, it did not allow us to establish the causality and temporality of the relationships that were observed. Selection bias may be significant if those of a poor people appear to have less participation. Fourth, there is risk of bias due to online data collection.

Study strengths are the novelty of surveying adult population in order to assess mental wellbeing at various career points. To the best understanding of the authors, there was no earlier sample of a general population study for mental health. The research was controlled enough to identify variations, as in comparable studies or settings. Further research is needed to find new findings, with larger and more diverse samples from populations with different health status.

## Conclusion

In this population-based analysis, we found that during this pandemic period mental well-being in the Bangladeshi adult population was worst in female, unemployed, business community occupational group and married person. Older people 50 years of age and over reported a greater day-to-day variation in their mental wellbeing ratings. The study will help to let practitioners and policymakers suggest where should apply the interventions to improve their mental health during and after this pandemic time.

## Data Availability

Click here for the data file http://individual.utoronto.ca/ahmed_3/index_files/data/data.html.

## Declarations

### Ethical approval

We conducted the study according to the guidelines laid down in the Declaration of Helsinki and all procedures involving human subjects were approved by the Institutional Review Board (IRB) of North South University (NSU-IRB 3572. Participants gave consent to participate in this study by accessing the online survey

### Consent to Publish

Not applicable.

### Availability of data

Click here for the data file http://individual.utoronto.ca/ahmed_3/index_files/data/data.html.

## Author’s contributions

MA and AH participated in study conception, design and coordination of the manuscript. GUA and HRK reviewed the manuscript and helped to draft the manuscript. AH also performed statistical analysis and helped to draft the manuscript. All authors approved the final manuscript.

## Acknowledgements

All the authors acknowledge the participants for providing us the information to conduct the study. We would also like to thank the three anonymous reviewers and the editor for insightful comments that improved the presentation and clarity of our manuscript.

## References

1. Rothan HA, Byrareddy SN. The epidemiology and pathogenesis of coronavirus disease (COVID-19) outbreak. J Autoimmun. 2020 Feb;102433. Available from: https://linkinghub.elsevier.com/retrieve/pii/S0896841120300469

2. Shereen MA, Khan S, Kazmi A, Bashir N, Siddique R. COVID-19 infection: Origin, transmission, and characteristics of human coronaviruses. J Adv Res. 2020 Jul; 24:24–91. Available from: https://linkinghub.elsevier.com/retrieve/pii/S2090123220300540

3. Cohen J, Kupferschmidt K. Strategies shift as coronavirus pandemic looms. Science (80-). 2020 Feb 28 [cited 2020 Apr 1];367(6481):962–3. Available from: https://www.sciencemag.org/lookup/doi/10.1126/science.367.6481.962

4. Ahorsu DK, Lin C-Y, Imani V, Saffari M, Griffiths MD, Pakpour AH. The Fear of COVID-19 Scale: Development and Initial Validation. Int J Ment Health Addict. 2020 Mar 27; Available from: https://doi.org/10.1007/s11469-020-00270-8

5. Cacioppo JT, Cacioppo S. Social Relationships and Health: The Toxic Effects of Perceived Social Isolation. Soc Personal Psychol Compass. 2014 Feb 1; 8(2):58–72. Available from: http://doi.wiley.com/10.1111/spc3.12087

6. Bhatti AB, Haq A ul. The Pathophysiology of Perceived Social Isolation: Effects on Health and Mortality. Cureus. 2017 Jan 24; Available from: http://www.cureus.com/articles/6213-the-pathophysiology-of-perceived-socialisolation-effects-on-health-and-mortality

7. Galea S, Merchant RM, Lurie N. The Mental Health Consequences of COVID-19 and Physical Distancing. JAMA Intern Med. 2020 Apr 10; Available from: https://jamanetwork.com/journals/jamainternalmedicine/fullarticle/2764404

8. Rohde N, D’Ambrosio C, Tang KK, Rao P. Estimating the Mental Health Effects of Social Isolation. Appl Res Qual Life. 2016 Sep 3; 11(3):853–69. Available from: http://link.springer.com/10.1007/s11482-015-9401-3

9. Mamun MA, Griffiths MD. Journal Pre-proof First COVID-19 suicide case in Bangladesh due to fear of COVID-19 and xenophobia: Possible suicide prevention strategies. Asian J Psychiatr. 2020;

10. Brooks SK, Webster RK, Smith LE, Woodland L, Wessely S, Greenberg N, et al. The psychological impact of quarantine and how to reduce it: rapid review of the evidence. Vol. 395, The Lancet. 2020. p. 912–20. Available from: https://doi.org/10.1016/

11. de Bruijn E-J, Antonides G. Determinants of financial worry and rumination. J Econ Psychol. 2020 Jan 1;76:102233. Available from: https://linkinghub.elsevier.com/retrieve/pii/S0167487019301679

12. Mckibbin W, Fernando R. Crawford School of Public Policy CAMA Centre for Applied Macroeconomic Analysis, The Brookings Institution Centre of Excellence in Population Ageing Research, The Global Macroeconomic Impacts of COVID-19: Seven Scenarios *. 2020; Available from: https://ssrn.com/abstract=3547729

13. Cacioppo JT, Hawkley LC. Social Isolation and Health, with an Emphasis on Underlying Mechanisms. Perspect Biol Med. 2003;46(3x):S39–52. Available from: https://doi.org/10.1353/pbm.2003.0063 https://muse.jhu.edu/article/44865

14. Anderson RM, Heesterbeek H, Klinkenberg D, Dã T, Hollingsworth irdre. How will country-based mitigation measures influence the course of the COVID-19 epidemic? 2020 Available from: https://doi.org/10.1016/

15. Day M. Covid-19: identifying and isolating asymptomatic people helped eliminate virus in Italian village. BMJ. 2020 Mar 23;368:m1165. Available from: http://group.bmj.com/group/rights-licensing/

16. Adhikari SP, Meng S, Wu Y-J, Mao Y-P, Ye R-X, Wang Q-Z, et al. Epidemiology, causes, clinical manifestation and diagnosis, prevention and control of coronavirus disease (COVID-19) during the early outbreak period: a scoping review. Infect Dis Poverty. 2020 Dec 17;9(1):29. Available from: https://doi.org/10.1186/s40249-020-00646-x

17. Cronbach LJ. Coefficient alpha and the internal structure of tests. Psychometrika. 1951 Sep;16(3):297–334.

18. Lai J, Ma S, Wang Y, Cai Z, Hu J, Wei N, et al. Factors Associated With Mental Health Outcomes Among Health Care Workers Exposed to Coronavirus Disease 2019. JAMA Netw Open. 2020 Mar 23;3(3):e203976. Available from: https://jamanetwork.com/journals/jamanetworkopen/fullarticle/2763229

19. Zhang Y, Ma ZF. Impact of the COVID-19 Pandemic on Mental Health and Quality of Life among Local Residents in Liaoning Province, China: A Cross-Sectional Study. Int J Environ Res Public Health. 2020 Mar 31;17(7):2381. Available from: https://jamanetwork.com/journals/jamanetworkopen/fullarticle/2763229

20. Gardner J, Oswald AJ. Money and mental wellbeing: A longitudinal study of medium-sized lottery wins. J Health Econ. 2007 Jan 1;26(1):49–60.

21. Godinic D, Obrenovic B, Khudaykulov A. Effects of Economic Uncertainty on Mental Health in the COVID-19 Pandemic Context: Social Identity Disturbance, Job Uncertainty and Psychological Well-Being Model. Int J Innov Econ Dev. 2019;6:61–74.

22. Pinho P de S, Araújo TM de. Associação entre sobrecarga doméstica e transtornos mentais comuns em mulheres. Rev Bras Epidemiol. 2012;15(3):560–72. Available from: http://www.scielo.br/scielo.php?script=sci_arttext&pid=S1415-790X2012000300010&lng=pt&tlng=pt

23. Shaw SCK. Hopelessness, helplessness and resilience: The importance of safeguarding our trainees’ mental wellbeing during the COVID-19 pandemic. Nurse Educ Pract. 2020 Mar 1 [cited 2020 May 15];44:102780. Available from: https://linkinghub.elsevier.com/retrieve/pii/S1471595320302791

24. Zhang SX, Wang Y, Rauch A, Wei F. Unprecedented disruption of lives and work: Health, distress and life satisfaction of working adults in China one month into the COVID-19 outbreak. Psychiatry Res. 2020 Jun 1;288:112958. Available from: https://linkinghub.elsevier.com/retrieve/pii/S0165178120306521

25. Ahmed MZ, Ahmed O, Aibao Z, Hanbin S, Siyu L, Ahmad A. Epidemic of COVID-19 in China and associated Psychological Problems. Asian J Psychiatr. 2020 Jun 1;51:102092. Available from: https://linkinghub.elsevier.com/retrieve/pii/S1876201820302033

26. Tweed E, Gounari X, Graham L. Mental wellbeing in prisoners in Scotland: an analysis of repeat cross-sectional surveys. Lancet. 2018 Nov 1;392:S11.

27. Hodiamont F, Allgar V, Currow DC, Johnson MJ. Mental wellbeing in bereaved carers: A Health Survey for England population study. BMJ Support Palliat Care. 2019 Sep 6;bmjspcare-2019–001957. Available from: http://spcare.bmj.com/lookup/doi/10.1136/bmjspcare-2019-001957

28. Koushede V, Lasgaard M, Hinrichsen C, Meilstrup C, Nielsen L, Rayce SB, et al. Measuring mental well-being in Denmark: Validation of the original and short version of the Warwick-Edinburgh mental well-being scale (WEMWBS and SWEMWBS) and cross-cultural comparison across four European settings. Psychiatry Res. 2019 Jan 1 [cited 2020 May 14];271:271–502. Available from: https://linkinghub.elsevier.com/retrieve/pii/S016517811831134X

29. Waqas A, Ahmad W, Haddad M, Taggart FM, Muhammad Z, Bukhari MH, et al. Measuring the well-being of health care professionals in the Punjab: a psychometric evaluation of the Warwick–Edinburgh Mental Well-being Scale in a Pakistani population. PeerJ. 2015 Oct 1;3(10):e1264. Available from: https://peerj.com/articles/1264

30. Fung S. Psychometric evaluation of the Warwick-Edinburgh Mental Well-being Scale (WEMWBS) with Chinese University Students. Health Qual Life Outcomes. 2019 Dec 14 17(1):46. Available from: https://doi.org/10.1186/s12955-019-1113-1

31. Santos JJA dos, Costa TA da, Guilherme JH, Silva WC da, Abentroth LRL, Krebs JA, et al. Adaptation and cross-cultural validation of the Brazilian version of the Warwick-Edinburgh mental well-being scale. Rev Assoc Med Bras. 2015 Jun 1 61(3):209–14. Available from: http://www.scielo.br/scielo.php?script=sci_arttext&pid=S0104-42302015000300209&lng=en&tlng=en

32. Smith ORF, Alves DE, Knapstad M, Haug E, Aarø LE. Measuring mental well-being in Norway: validation of the Warwick-Edinburgh Mental Well-being Scale (WEMWBS). BMC Psychiatry. 2017 Dec 12;17(1):182. Available from: http://bmcpsychiatry.biomedcentral.com/articles/10.1186/s12888-017-1343-x

33. Murray MA, Cardwell C, Donnelly M. GPs’ mental wellbeing and psychological resources: a cross-sectional survey. Br J Gen Pract 2017; 67(661): e547–e554. DOI: https://doi.org/10.3399/bjgp17X691709

